# From Evidence to Intervention: A Systematic, Multi-Stakeholder Approach to Developing and Prioritizing Diagnostic Excellence Interventions Across a 14-Site Hospital Medicine Collaborative

**DOI:** 10.64898/2026.10.22.26366208

**Authors:** Jeffrey L. Schnipper, Katie E. Raffel, James D. Harrison, Robert S. Young, Andrew P.J. Olson, Gopi J Astik, Jeffrey A. Gold, Abhishek Goyal, Peter Barish, Anuj K. Dalal, Sumant R. Ranji, Allison R. Bond, Tiffany Lee, Andrew D. Auerbach, the ADEPT Study Group

## Abstract

**Background:** Diagnostic errors are common and represent a leading cause of preventable harm in hospitalized medical patients. Despite growing recognition of this problem, the field lacks a systematic, evidence-based approach for developing, adapting, and selecting diagnostic excellence interventions at scale across multiple institutions.

**Methods:** Using a structured, multi-phase process, the Achieving Diagnostic Excellence through Prevention and Teamwork (ADEPT) collaborative, a 14-site AHRQ-funded hospital medicine network: (1) conducted an environmental scan of existing diagnostic safety practices across sites; (2) constructed a conceptual framework organizing potential interventions by target (patient, provider/team, health system) and safety paradigm (Safety I and Safety II); (3) convened multidisciplinary working groups to develop detailed proposals for six intervention programs; (4) applied a pre-specified multi-criteria rubric to facilitate group deliberation, and (5) voted to select a single high-priority intervention for collaborative launch.

**Results:** Six interventions were developed, selected, or adapted from prior work, spanning all cells of the target and safety paradigm framework. Finalized proposals were then shared at two separate ADEPT meetings – the first to solicit feedback on interventions and to coordinate activities across groups, and the second where a formal ranked-choice vote was held. This process resulted in selection of the Diagnostic Cross-Check (DXC) — a structured, peer second-opinion program triggered by rapid response team (RRT) events — as the highest priority intervention for collaborative-wide launch. The remaining interventions were designated as components of an ADEPT toolbox for future adaptation, adoption, and evaluation.

**Discussion:** The ADEPT collaborative’s systematic, evidence-informed approach to diagnostic intervention development offers a replicable model for other multi-site quality improvement networks. The resulting portfolio of interventions provides a roadmap for the field of inpatient diagnostic excellence.

## Background

Diagnostic errors — defined as a missed opportunity to make a timely or correct diagnosis, based on the available evidence, regardless of patient harm^1,2^ — affect approximately 5% of outpatient encounters and remain one of the most consequential and under-addressed sources of preventable patient harm.^3-5^ In the inpatient setting, it is estimated that 7% of general medicine patients have experienced a harmful diagnostic error and 6% have experienced a preventable one.^6^ Among medicine inpatients who died or were transferred to the intensive care unit, 23% experienced a diagnostic error.^7^ Errors contributed to temporary harm, permanent harm or death in 18% of all evaluated patients, and among patients who died, the diagnostic error contributed to the patient’s death in 7%.^7^ The most common underlying causes of inpatient diagnostic errors lie in clinical assessment and issues with test ordering, performance, and interpretation.

Despite this burden, the diagnostic safety field has lagged behind other patient safety domains in developing, testing, and scaling evidence-based interventions. Existing efforts have largely been single site, focused on outpatient or emergency department settings or on narrow types of errors (e.g., radiology), and have not addressed the full complexity of the diagnostic process across patient, provider, and health system levels.^8^

The Achieving Diagnostic Excellence through Prevention and Teamwork (ADEPT) collaborative was established to address this gap. Funded by the Agency for Healthcare Research and Quality (AHRQ), ADEPT comprises 14 hospital medicine programs across the United States and aims to advance the science of diagnostic safety through multi-site surveillance, an analysis of underlying causes, benchmarking, and the development and evaluation of targeted interventions.

A critical and previously undescribed challenge for the collaborative was how to systematically move from empirical findings about the causes of diagnostic error to a prioritized, implementable set of interventions. The diagnostic process is complex, multi-level, and contextually variable — meaning no single intervention is likely to be sufficient, yet resources constrain the number that can be pursued concurrently.

In this paper, we describe ADEPT’s approach to intervention program selection (with interventions either developed *de novo* or adapted from prior work): the conceptual framework we employed, the process by which working groups developed detailed proposals, the criteria and deliberative mechanism used to select a priority intervention, and the resulting portfolio of future interventions. We offer this account both as a contribution to the diagnostic excellence literature and as a methodological model for other multi-site quality improvement collaboratives.

## Methods

### Setting and collaborative structure

ADEPT is a 14-site hospital medicine collaborative funded by AHRQ (Grant No. 1R18HS029366).^9^ Participating sites are members of the Hospital Medicine Reengineering Network (HOMERuN), a national research collaborative,^10^ and span academic medical centers, community hospitals, and safety-net institutions across the United States. The collaborative conducts prospective, structured case reviews of rapid response team (RRT) activations, ICU transfers, and inpatient deaths on medicine services to identify diagnostic errors using a modified version of the SaferDx tool, as in the previously conducted Utility of Predictive Systems in Diagnostic Errors (UPSIDE) study ^11,12^ and characterizes their underlying causes using a modified version of the Diagnostic Error Evaluation and Research (DEER) taxonomy.^13^ At the time of intervention program development/adaptation/selection (hereafter referred to simply as development), 3467 cases had been adjudicated across all study sites. As with UPSIDE, the most common underlying causes of diagnostic errors were issues with clinical assessment (including failure or delay in considering the diagnosis and suboptimal weighing or prioritizing diagnoses), test ordering and interpretation, suboptimal weighing of a piece of history data, and failure or delay in acting or following up on a test result.

### Phase 1: Environmental scan

Prior to intervention program development (January – April, 2024), the ADEPT team conducted a systematic environmental scan of existing diagnostic safety practices across participating sites.^14^ Site leads documented current programs — including morbidity and mortality conferences, case review committees, event reporting systems, EHR-based triggers, and provider feedback mechanisms — using a structured template. This scan served to identify gaps in existing infrastructure, avoid duplication of existing efforts, and contextualize the intervention landscape at each institution.

### Phase 2: Conceptual framework development

The collaborative then applied a 3×2 theory-based framework to our environmental scan and literature review as a way to organize the space of potential diagnostic safety interventions (**Table 1**). The framework dimensions were: 1) The target of the intervention: patient, clinician/team, or health system, and 2) The safety paradigm: Safety I (preventing failures and reducing error rates) versus Safety II (building resilience, adaptability, and the conditions under which clinicians succeed).^15^ Application of the framework was also informed by ADEPT’s empirical findings — specifically, the predominance of provider assessment and clinical reasoning as common cause contributors — and by existing conceptual models of diagnostic process improvement.^3,5,16^ Each cell of the matrix was assigned a dedicated working group comprising investigators and clinical leads from multiple participating sites.

**Table 1.** ADEPT Intervention Framework.

|  | Safety I (Prevent Failure) | Safety II (Build Resilience) |
| --- | --- | --- |
| **Patient** | Communication & Patient Activation (SHARE-DX Framework + Patient Diagnostic Feedback Tool) | ↑ Same interventions span both paradigms |
| **Provider / Clinician** | Self-directed Reflection & Feedback (ADEPT-Reflect Tool) | Diagnostic Time-Out (DTO)<br>EHR Best Practices (Epic user optimization)<br>LLM tools for diagnosis |
| **Health System** | Embedding Diagnostic Error Review into Safety/Quality Infrastructure | Second Opinion Program →<br>Diagnostic Cross-Check (DXC) ★ |

ADEPT: Achieving Diagnostic Excellence through Prevention and Teamwork; EHR: Electronic Health Record; LLM: large language model

### Phase 3: Working group proposal development

Between January and March 2025, each working group, consisting of 5-20 members from the wider ADEPT study team, developed a detailed intervention proposal using a standardized template. Proposals addressed: (1) the specific diagnostic gaps targeted, grounded in ADEPT case review findings (i.e., the most common underlying causes of errors); (2) safety behavior target (i.e., in which cell of Table 1 it belonged); (3) background and description of the intervention, including intervention design and components; (4) measurement approach; (5) references and evidence; (6) potential implementation approaches, from low resource intensity to high resource intensity; (7) training needs; and (8) an explicit assessment of feasibility, measurability, alignment with each site’s organizational priorities, and potential impact. Working groups conducted literature reviews, reviewed environmental scan data, and engaged in iterative refinement through 3-5 virtual meetings. A summary of all proposals was compiled and circulated to the full collaborative in advance of a structured review session.

### Phase 4: Deliberative review and formal prioritization

In March 2025, we sent a summary presentation of all interventions to all ADEPT stakeholders, including Chief Quality Officers (CQOs), Advisory Committees, Diagnostic Excellence Consultants, Site leads, and Adjudicators for asynchronous review. We conducted separate meetings with CQOs, Advisors, and Consultants for facilitated discussion with study leadership. On April 3, 2025, an All-Hands meeting convened site leads and adjudicators for structured deliberation, during which time each working group presented their proposal (approximately 10 minutes), followed by facilitated discussion addressing questions, suggesting refinements, and considering initial feasibility assessment across sites.

During this final meeting, all proposals were evaluated against a pre-specified seven-criteria prioritization rubric (**Table 2**).

**Table 2.** Intervention Prioritization Criteria.

| **Criterion** | **Operationalization** |
| --- | --- |
| Feasibility | -Within hospital medicine control<br>-Implementable within 12 months across participating sites |
| Measurability | -Impact assessable through existing data infrastructure or low-burden surveys |
| Organizational alignment | -Consistent with institutional quality/safety priorities at ADEPT sites |
| Stakeholder support | -Site leadership willing to give it resources and support<br>-Hospital medicine group willing to put effort into implementation |
| Clinical impact | -Targets the most prevalent underlying cause domains identified in ADEPT case reviews<br>-Impact will be meaningful |
| Evidence base | -Supported by existing literature or, where novel, no evidence of harm |
| Multi-site adaptability | -Sufficiently flexible for implementation across diverse hospital contexts |

ADEPT: Achieving Diagnostic Excellence through Prevention and Teamwork

Following the meeting, a formal ranked-choice ballot was distributed to all ADEPT stakeholder groups to identify the intervention to be prioritized for collaborative-wide pilot launch in May–June 2025. The remaining interventions were designated as the ‘ADEPT toolbox’ — resources available for local adoption and future collaborative study.

## Results

### Environmental scan findings

The environmental scan revealed substantial variation in existing diagnostic safety infrastructure across the 14 sites. Most sites had at least one active case review mechanism (M&M conferences, hospital medicine mortality review, and/or hospital-level safety event reporting), but these were not systematically focused on diagnostic error identification. Few sites had structured programs specifically targeting diagnostic reasoning, patient engagement in the diagnostic process, or provider feedback linked to diagnostic outcomes. Most interventions were reactive in nature (e.g., reviewing cases where an adverse event occurred) rather than proactive (e.g., identifying cases in real-time to prevent errors from occurring or mitigate their effects). Clinicians reported that they desired greater support in the diagnostic process (especially when cognitive load is high), a supportive culture for feedback, and interventions that acknowledged context of care and resource constraints. Lastly, site leads agreed that successful programs need to focus on both systems issues and clinician performance.^14^

### Intervention program proposals

Six intervention programs were developed across the framework (see **Appendix 1** for each of the proposals).

### Patient interventions

The Patient Communication and Activation working group and HOMERuN Patient & Family Advisory Council developed ‘Bringing Doctors Back to the Bedside,’ comprising two complementary tools: (1) the Patient and Family Diagnostic Feedback Tool — a structured electronic survey administered within the first 72 hours of hospitalization, asking patients to reflect on their diagnosis and flag perceived discrepancies^17-19^ — and (2) the SHARE-DX Framework, a practical bedside framework for integrating patient and family engagement into the diagnostic process for hospitalized patients. The mnemonic for SHARE-DX is Set the Stage & Summarize, Hypothesize together; Align with Lived Experience; Review for Red flags, Risk & Roadmap; Educate & Engage (see **Appendix 2**). SHARE-DX translates evidence and national guidance on diagnostic safety, diagnostic reasoning, and patient-centered communication into structured bedside behaviors. SHARE-DX builds on prior work, most notably the Uncertainty Communication Toolkit.^20^ This intervention was classified as spanning both Safety I (error prevention through patient-flagged discrepancies) and Safety II (building diagnostic resilience through co-production of a diagnostic plan).

### Provider interventions

Four provider-facing interventions were developed. The Provider Self-Reflection and Feedback program (Safety I) proposed deploying the ADEPT-Reflect tool — a structured, confidential reflection instrument modelled on the Diagnosis Learning Cycle^21^ and CalibrateDx framework^1,22^ — to clinicians following trigger events (RRT, ICU escalation), with optional coaching and anonymized peer sharing of learnings based on structured case review (see **Appendix 2** for the tool).

The EHR Best Practices program (Safety II) proposed training clinicians in optimal Epic use for diagnostic purposes, using a train-the-trainer model developed by Miller et al. at OHSU^23^ and an adapted library of short instructional videos,^24^ with optional 1:1 coaching, especially if evidence of suboptimal EHR use based on use data.

The Diagnostic Time-Out (DTO) intervention (Safety II), adapted from prior work by Dalal et al.,^17^ aimed to integrate a structured, evidence-based pause into clinical workflow at trigger points (e.g., RRT, handoffs, clinical deterioration), prompting teams to revisit the working diagnosis, consider cognitive debiasing, and document the DTO via an EHR structured template.

Lastly, while not formally proposed, LLM Tools for Diagnosis (Safety II) developed concrete guidance on when and how to use accessible LLMs to generate differential diagnoses based on patient presentations, including what models to use, how to provide patient data to the LLM in a HIPPA-compliant manner, and how to write a prompt to optimize the information delivered. This work also informed the development of a manuscript on the “5 rights” of LLM-assisted diagnosis in the inpatient setting.^25^

### Health system interventions

Two system-level interventions were developed. The Embedding Diagnostic Safety into Quality/Safety Infrastructure program (Safety I) proposed a hub-and-spoke model in which ADEPT teams would build diagnostic error case review into existing hospital safety and quality committees, with a central diagnostic excellence team forming the institutional hub of experts needed to coordinate diagnostic excellence improvement efforts across many domains (such as a case review framework, coordination with incident reporting and root-cause programs, and integration with provider coaching programs). This group also laid out needs for legal peer-review protections, escalation pathways for system-level findings, and structures for cross-departmental spread. Because this particular domain had substantial overlap with the scientific and operational aims of ADEPT, the group chose to treat this particular intervention development as a separate task for ADEPT, and it was not included in later refinement or voting processes.

The Second Opinion Program (Safety II) proposed formalizing peer consultation for perceived diagnostic uncertainty within hospital medicine, ranging from lower-intensity dyad/pod structures to a designated second-opinion clinician-of-the-day who proactively contacts on-service teams. This intervention subsequently evolved into the Diagnostic Cross-Check (DXC), described below.

### Prioritization process and outcome

#### Final intervention selection

Following the April 2025 All-Hands deliberation, a formal collaborative vote identified the Diagnostic Cross-Check (DXC) as the priority intervention for coordinated launch, based on 32 stakeholder responses, with 14 initial first-choice votes, and 19 votes by the end of the ranked-choice (instant run-off) voting process. The diagnostic time-out came in second place with 8 initial first-choice votes.

DXC was favored on several grounds. It directly targeted the predominant underlying cause domain identified in ADEPT reviews (provider assessment and clinical reasoning), with a structured peer consultation triggered by RRT activation — a sentinel event already embedded in clinical workflow across all sites. It had the strongest existing evidence base within the collaborative, demonstrating a 40% relative reduction in adverse events when conducted in the emergency department setting,^26^ clearest measurement pathways using short provider surveys, and was adaptable to diverse site contexts without requiring new infrastructure. Its Safety II framing — building a culture of normalized diagnostic uncertainty and peer support — was viewed as particularly consonant with the collaborative’s goals. The DXC was designed to be proactive, identifying patients who might be in the early stages of a diagnostic error, where prompt changes in the diagnostic plan could improve care for that patient. In that sense, its timing and structure contained some elements of the Diagnostic Time-Out. At the same time, the DXC also contained some elements of the Feedback intervention, especially in cases where it became clear that a diagnostic error had in fact occurred, with a goal of changing the way clinicians care for future patients. Lastly, over time, cross-checkers increasingly incorporated the use of LLMs in their processes, e.g., in cases of diagnostic mystery where a broad differential diagnosis was desired by the clinician. In other words, the DXC evolved to absorb elements of several other proposed interventions.

While Embedding Diagnostic Safety into Quality/Safety Infrastructure was not chosen, it is an essential element of the ADEPT study (and any program designed to improve diagnostic performance). Thus, all sites also agreed to incorporate this element into usual care as they work to sustain diagnostic excellence well beyond the period of the ADEPT study.

The DXC was pilot tested in the summer of 2025, with very promising results: based on brief surveys of attendings who received them, in 1/6 of cases, it led to a change in the diagnostic care plan for that patient, about 40% of clinicians said it would change the way they care for future patients, and 99% said they would be willing to do another cross-check in the future.^2^ Based on those findings, the DXC was formally launched at all ADEPT sites on October 6, 2025.

## Discussion

The ADEPT collaborative’s experience offers several insights relevant to the broader field of diagnostic excellence and multi-site quality improvement.

By organizing potential interventions along two orthogonal dimensions — target level and safety paradigm — the collaborative was able to ensure comprehensive coverage of the diagnostic problem space, avoid implicit bias toward any single class of solution, and communicate the intervention portfolio to diverse stakeholders in a structured way. The Safety I / Safety II distinction proved particularly generative: it surfaced the observation that most existing diagnostic safety efforts are Safety I in orientation (error detection and reduction), while the most promising opportunities for durable culture change may lie in Safety II approaches that strengthen the conditions for diagnostic success. On the other hand, some interventions clearly contained both Safety I and Safety II components, especially the Discharge Time-Out and SHARE-DX framework. For example, the Diagnostic Time-Out can facilitate diagnostic error detection and retrospective case-based learning as well as promoting resilience in the diagnostic decision-making.

The Delphi-like process we used appropriately balanced structure and rigor in assessment of quality improvement interventions, while being deeply collaborative and front-line clinician-focused. Our ADEPT prioritization process shares features with formal Delphi methodology — iterative expert input, structured criteria, and formal convergence — while adapting these to the pragmatic nature of the collaborative. The use of a pre-specified rubric was essential: it structured deliberation, made trade-offs explicit, and provided a defensible basis for the final decision. We suggest that this approach is generalizable to other multi-site QI networks facing similar challenges of moving from evidence to prioritized action.

DXC’s selection reflects a broader principle: when resources are limited, priority should be given to interventions with the strongest existing evidence, clearest trigger structures, and greatest site-level adaptability. The voting process itself also generated buy-in among site leads and other ADEPT participants in that they felt investment in the intervention that was chosen.

The ADEPT toolbox resulting from our work – even those not adopted in the short term by ADEPT - can provide a methodologic approach and potential intervention roadmap for the field of diagnostic excellence in hospital settings. Crucially, the collaborative process produced not just one intervention but a systematically developed portfolio. Each of the five non-priority interventions represents a distinct, evidence-informed approach to a different mechanism of diagnostic failure. Publishing these proposals — as this paper does collectively, and soon as individual tool tutorials — contributes to a nascent but growing library of testable diagnostic excellence interventions. We hope this portfolio accelerates health delivery and implementation science in this domain.

Several limitations warrant acknowledgment. First, our framework was focused on hospital medicine and may not generalize to other clinical settings. Second, the working group proposals, while grounded in ADEPT data and existing literature, have not all undergone formal pilot testing; feasibility and effectiveness data will be reported separately. Third, the prioritization process, although structured, involved elements of group dynamics and institutional influence that cannot be fully eliminated from any collaborative decision-making process. Finally, the Safety I / Safety II dichotomy, while conceptually useful, oversimplifies a complex continuum of safety approaches. Indeed, as noted above, several of these interventions could be said to incorporate elements of both Safety I and Safety II.

## Conclusion

The ADEPT collaborative has developed a systematic, framework-driven approach to diagnostic excellence intervention development that may serve as a model for the field. By grounding proposals in multi-site empirical data, organizing them within a coherent conceptual framework, and applying a structured prioritization process, the collaborative has produced both a priority intervention (DXC) and a portfolio of future candidates. Together, these represent a meaningful step toward a more intentional, evidence-based approach to reducing diagnostic harm in hospital medicine.

## Supporting information

Appendix 2 - Intervention Tools

Appendix 1 - Intervention Templates

## Data Availability

All data produced in the present study are available upon reasonable request to the authors

https://hospitalinnovate.org/projects/adept-study/

## Acknowledgements

We would like to thank all the hospital leaders, including Chief Quality Officers, Hospital Medicine Division and Section Chiefs, and quality and safety leadership; ADEPT adjudicators; Diagnostic Cross-checkers; and front-line clinicians who participated in the ADEPT study for their dedication to patient care and willingness to participate in this study.

## Notes

Declaration of Interest Statement: The authors declare no conflicts of interest relevant to the work.

This work was supported by the Agency for Healthcare Research and Quality (Grant No. 1R18HS029366). The funder had no role in study design, data collection, analysis, or the decision to publish.

**Transparency and ethics** Data availability: Intervention proposal templates are available as supplementary materials, and study data will be made freely available to support implementation by other organizations upon request.

### Competing Interest Statement

The authors have declared no competing interest.

### Clinical Protocols

https://pubmed.ncbi.nlm.nih.gov/37888951/

