## Appendix 2 - Intervention Tools for "From Evidence to Intervention: A Systematic, Multi-Stakeholder Approach to Developing and Prioritizing Diagnostic Excellence Interventions Across a 14-Site Hospital Medicine Collaborative"

**ADEPT Intervention Tools** **Page**

1. **ADEPT-Reflect Tool** **2**
2. **SHARE-Dx Tool** **5**
3. **Diagnostic Cross-Check Tool** **7**

### ADEPT-Reflect Tool

*Note – This is a preliminary design for this tool, and the tool could be adapted and refined after initial use.*

Reflection is fundamental for improving performance. The purpose of this tool is to foster a brief reflection among members of a clinical hospital medicine team after a care transition, such as transfer to the Intensive Care Unit. This can also be used to allow reflection after care is transitioned to another care team, such as at the end of a week of service, and the diagnosis changed.

This entire exercise should take no more than 10 minutes, although you are encouraged to follow up in more detail regarding learnings you may have identified. This tool is for individual and team learning.

*Please avoid putting any PHI into this tool.*

How to complete the ADEPT-Reflect Tool:

1. Identify a case for which there was change in diagnosis after a transition in care or a transfer to a higher level of care.
    a. Examples include ICU transfers, transition to another team/team member, transfer to another hospital, etc.
2. Set aside 10 minutes to complete this tool. This can be done as an individual, but it is best done as a care team.
3. Try to complete 2 of these weekly.
4. The ADEPT team will capture the learnings you identify (if you clicked “OK to share”) and will share these in a structured, deidentified way with your group.

Select which type of case you are reviewing:

[ ] Transfer to higher level of care

[ ] Change in diagnosis after a transition of care

*If “Transfer to higher level of care” is selected:*

1. Describe briefly (3-4 sentences) why this patient was transferred to the higher level of care.
2. Were you surprised by this?
    a. Why or why not?

3. What was your main working diagnosis (or few working diagnoses) prior to the transfer?
    a. Did new working diagnoses arise during the process of the transfer?
    b. Did your prioritization of differential diagnosis items change as part of the transfer?
4. Did any new diagnostic information arise during the process of the transfer?
    a. If yes, what? How did this arise?
5. Think about your or your team's diagnostic processes leading up to the transfer to a higher level of care. We know that the following areas are particularly challenging with respect to the diagnostic process in hospital medicine. Which of these areas (one or more than one) would you like to reflect upon further?
    a. How decisions were made around laboratory and imaging tests
    b. How patients were monitored and/or how diagnostic information was followed up
    c. How clinical reasoning occurred among individuals and teams, including synthesis and weighing/prioritization of information, consideration of potential diagnoses, over- or under-reliance on tests or consultations, and other issues.
6. Based on the area(s) you selected, describe how something in this area arose during the transfer to a higher level of care that you might consider doing differently next time. What is it? Why?
7. There are also things that are always done well in a case, even when a clinical outcome is challenging. What is something that went well in this case?
8. What specific learnings will you carry forward from this case? Is it ok to share this with the group?

*If "Change in diagnosis after a transition of care" is selected:*

1. Describe briefly (3-4 sentences) what diagnos(es) changed.
2. Were you surprised by this?
    a. Why or why not?
3. What was your main working diagnosis (or few working diagnoses) prior to this change?
4. Did any new diagnostic information arise?
    a. If yes, what? How did this arise?
5. Think about your or your team's diagnostic processes leading up to this diagnostic change. We know that the following areas are particularly challenging with respect to the diagnostic process in hospital medicine. Which of these areas (one or more than one) would you like to reflect upon further?
    a. How decisions were made around laboratory and imaging tests
    b. How patients were monitored and/or how diagnostic information was followed up

    c. How clinical reasoning occurred among individuals and teams, including synthesis and weighing/prioritization of information, consideration of potential diagnoses, over- or under-reliance on tests or consultations, and other issues.

6. Based on the area(s) you selected, describe how something in this area arose during the change in diagnosis that you might consider doing differently next time. What is it? Why?
7. There are also things that are always done well in a case, even when a clinical outcome is challenging. What is something that went well in this case?
8. What specific learnings will you carry forward from this case? Is it ok to share this with the group?

### SHARE-Dx Tool: A Framework for Diagnostic Partnership

| Domain | Purpose | Core Behaviors / Techniques | Example Scripted Language |
|---|---|---|---|
| S – Set the Stage & Summarize | Establish partnership, invite the patient's story, and empower them (and their care partners) to share or correct the clinician's understanding. This step builds on the foundational practice of uninterrupted listening—such as the "Sixty Seconds to Improve Diagnostic Safety" approach used in the outpatient setting—by integrating what was heard into a shared summary and diagnostic dialogue. | • Sit at eye level, introduce roles, and frame diagnosis as a shared process.<br>• Uninterrupted listening to the patient's story.<br>• Normalize correction by explaining this is done with every patient.<br>• Summarize what is known so far and explicitly invite "edits" to your understanding. | "You know your body and story best. I want you to feel comfortable correcting me if something sounds off. Here's what we've gathered so far—what feels most important to you, or what might I be missing?" |
| H – Hypothesize Together | Share diagnostic reasoning transparently and test hypotheses collaboratively. | • Confirm the key facts of the patient's story and problem(s).<br>• Name 2–3 working diagnoses and describe the reasoning for each ("the why").<br>• Explain what findings would make each diagnosis more or less likely.<br>• Invite patient or family input on which explanations seem plausible. | "Here is what I am prioritizing as the key features. Based on what you've told me, the main possibilities are [A], [B], and [C]. Each has certain features we look for [specific disease characteristics], which makes [diagnosis] seem most likely." |
| A – Align With the Lived Experience | Elicit how proposed explanations fit—or conflict—with the patient's story, values, and priorities. | • Ask whether diagnostic explanations align with the patient's experience.<br>• Explore discrepancies or contextual factors that influence interpretation.<br>• Integrate patient goals and concerns into next diagnostic steps. | "Does this explanation fit with what you've been feeling or what concerns you most?" |
| R – Red Flags, Risk & Roadmap | Engage patients and families as partners in ongoing surveillance by sharing what to expect and when to worry. | • Address the degree of diagnostic certainty.<br>• Explain the working diagnosis, treatment plan, and expected recovery course.<br>• Identify specific red flags, medication side effects, or patterns that | "While we feel [diagnosis] is very likely, we aren't completely certain. Here's what we expect over the next few days [roadmap]. If instead you notice [specific red flags], please let us know—it might mean we need to rethink the diagnosis." |

| | | | |
|---|---|---|---|
| | | should prompt re-evaluation.<br>• Clarify when to re-contact the team or escalate concern. | |
| E – Educate & Engage | Confirm shared understanding, ensure teach-back, and close the loop on communication. | • Begin by inviting one last concern or intuition.<br>• Use teach-back ("Can you tell me how you'll explain this to your family?").<br>• Reinforce the patient's key questions about diagnosis, next steps, and warning signs. | "Just so I know I explained this clearly, can you tell me what you understand about the diagnosis and plan? Is there anything that still feels off?" |

### Diagnostic Cross-Check Tool

1. What is your main working diagnosis for the reason for the RRT?
2. Is there anything about this patient's case that "doesn't fit" your working diagnosis?
3. What alternative diagnoses are you considering?
4. Based on the clinical picture before and immediately after the RRT, consider this checklist:
   - [ ] What additional chart review or history from patients or family members you should gather or reassess?
   - [ ] What aspects of the patient's physical examination should you gather or reassess?
   - [ ] Which tests (labs, xrays) you should order, follow-up on, or reinterpret?
   - [ ] Which consultations or additional team members you think would be helpful?
5. Based on this review, what are your next steps for this patient?
6. What aspects of the diagnostic process you think went well? What lessons do you want to share with others?
