## Appendix 1 - Intervention Templates for "From Evidence to Intervention: A Systematic, Multi-Stakeholder Approach to Developing and Prioritizing Diagnostic Excellence Interventions Across a 14-Site Hospital Medicine Collaborative"

**Intervention Templates** **Page**


| | |
|---|---|
| 1. Bringing Doctors Back to the Bedside: Engaging Patients to Reduce Diagnostic Errors. | 2 |
| 2. Provider Self-Reflection and Feedback | 8 |
| 3. EHR Training and Best Practices | 14 |
| 4. Diagnostic Time-Out | 18 |
| 5. Embedding Diagnostic Safety into Safety/Quality infrastructure | 22 |
| 6. Second Opinion Programs to Improve Diagnostic Performance | 28 |

### ADEPT Intervention Proposal

#### Bringing Doctors Back to the Bedside: Engaging Patients to Reduce Diagnostic Errors.

1. **Intervention descriptions:**

   Our Patient Communication and Activation Committee developed *Bringing Doctors Back to the Bedside: Engaging Patients to Reduce Diagnostic Errors.*

   This intervention consists of two complimentary tools to be used concurrently:
   1. The Patient and Family Diagnostic Feedback Tool, modeled after *Be the Expert on You* and adapted to the inpatient setting for use by patients and families.
   2. SHARE, a scripted, structured communication instrument designed to support physicians inviting patients and caregivers to engage in collaborative diagnosis.

2. **Priority ADEPT diagnostic gaps addressed with examples from our data:**

   **Clinical Reasoning/Assessment**
   At a fundamental level, both the Patient and Family Diagnostic Feedback and SHARE Tools seek to improve diagnostic accuracy by enabling patients and families to ensure that critical history and symptoms are incorporated into the provider's diagnostic decision-making process.

   **History and Physical Exam**
   This intervention provides patients with the time and space to reflect on their phenomenology (including physical exam symptoms) to deliver accurate diagnostic data to providers. It does so through the asynchronous use of the Patient and Family Diagnostic Feedback Tool and a structured, iterative discussion between the patient and provider, implemented by the SHARE script and *60 Seconds to Improve Diagnostic Safety.*

   **Healthcare Team Dynamics**
   In the 2015 *National Academy of Medicine Report*, *Improving Diagnosis in Health Care*, the first listed goal is to facilitate more effective teamwork in the diagnostic process. The *Bringing Doctors Back to the Bedside* intervention accomplishes this by integrating patients, families, and healthcare providers into the diagnostic process as a team.

3. **Safety behavior target**
   Our intervention is both a Safety I and II intervention:
   **Safety I:**
   Engaging patients and families can help prevent failures and errors. For instance, close communication and patient/family involvement can alert providers when a diagnosis may not be correct or to the need for an escalation of care in acute, life-threatening illnesses.

**Safety II:**
Improved patient and family communication, combined with enhanced metacognition techniques, strengthens the robustness, resilience, and adaptability of the overall diagnostic process.

4. **Background and description of the intervention:**

**The Patient and Family Diagnostic Feedback Tool:** A structured, asynchronous patient feedback tool can provide an additional layer of diagnostic safety.

Modeled on the Be the Expert on You Tool , this tool is administered within the first 72 hours of hospitalization via electronic survey (e.g., REDCap, Qualtrics), this tool would ask key questions:

1. What is your understanding of the main reason for your hospitalization?
2. What diagnosis has the medical team provided and is currently treating?
3. Do you feel like they have the right diagnosis?
4. Do you feel like you are improving, worsening, or staying the same?
5. What parts of your medical history do you think the medical team may be overlooking or not fully aware of?

These responses could serve as real-time diagnostic red flags, triggering a "diagnostic timeout" when significant discrepancies are identified. These questions could be used by patients daily to help calibrate their own thinking while participating in the diagnostic process with the provider. These questions were curated with patients and families.

**The physician-facing, SHARE-DX Framework for Collaborative Diagnosis (as requested by the PFAC):** To maximize the effectiveness of patient engagement, physicians need structured communication techniques. The SHARE-DX framework provides a systematic approach to integrating patient input into diagnostic reasoning:

**1. Summarize & Set the Stage**
a. "I'm going to summarize my understanding of your health so far and what brought you here. Please correct anything I have wrong or fill in any gaps."
b. Ensures the right problem is being diagnosed by refining key historical details (e.g., timeline, symptom evolution).
c. 60 Seconds to Improve Diagnostic Safety (AHRQ). Listen to the patient for 60 seconds without interruption.

**2. Hypothesize Together**
a. "This sounds most like [Dx] based on [symptoms, findings, test results]. Is this what you are experiencing?"
b. "There are a few possibilities based on my assessment: [outline reasoning]. The best approach is to rule out 'X' while managing for 'Y.' Does that sound reasonable?"
c. Engages the patient in the clinical reasoning process by articulating and testing diagnostic hypotheses.

**3. Align with the Patient's Lived Experience**
a. "This sounds like 'X'. X typically presents with a, b, c symptoms over y time course. Patients with prior history of 'Z' are at greatest risk. The data typically shows… Does that sound right to you?"
b. Checks if the diagnosis aligns with the patient's lived experience and invites them to refine or challenge the proposed illness script.

**4. Review for Red Flags, Risk and Roadmap**
a. "Are there any symptoms or experiences that don't fit with this explanation?"
b. "Sometimes, doctors make assumptions that aren't quite right. Is there anything I've said that doesn't seem right to you?"
c. Encourages patients to highlight inconsistencies that may warrant revisiting the differential diagnosis.

**5. Educate and Engage**
a. "Have I missed anything that feels important to you?"
b. "What am I missing or not accounting for?"
c. Activates System 2 reasoning by prompting the physician to slow down and consider alternative explanations.
d. Patient Teach Back to confirm understanding

*Potential Measurement Strategies*:
The AHRQ IDEAS project promotes the "Be the Expert on You" and "60 Seconds to Improve Diagnostic Safety," and recommends the Patient Exit Survey and the 60 Seconds to Improve Diagnostic Safety Provider Feedback Survey as evaluations for these two tools. These can be adapted and used for our patients using the new intervention.

https://www.ahrq.gov/diagnostic-safety/tools/engaging-patients-improve.html

Survey patients and family regarding provider compliance with the asynchronous tool and the script in their interactions with providers. DE rates once intervention implemented (factor analysis).

#### 5. References/Evidence:

The Toolkit for Engaging Patients to Improve Diagnostic Safety- https://www.ahrq.gov/diagnostic-safety/tools/engaging-patients-improve.html

As proof of concept for the Patient and Family Diagnostic Feedback Tool and the SHARE Framework for Collaborative Diagnosis our group found "OurDx." This instrument was operationalized as an online pre-visit survey which scripted similar questions/domains as our proposed instrument:

**Table 2.** OurDX domains, items, rationale, and patient completion rates

| OurDX domain | Patient contribution | OurDX item | Rationale (goal of item) | Site 1 completion rate N = 7075 | Site 2 completion rate N = 460 |
|---|---|---|---|---|---|
| Share what matters most to you (priorities) | Patient/family visit priorities (free text) | What are the most important things you would like to talk about at your visit? | Help patients to feel heard;[17,43] align patient-provider visit agendas.[46] | 5906 (83.5%) | 276 (60.0%) |
| Tell us how you have been (recent medical history) | Medical history (checkboxes) | Over the last 6 months (or since your last visit in this clinic), have you had any of the following? (New/worsening symptoms; chronic conditions; tests or procedures; visits to UC or ED or another hospital or HCP for the same problem; medication changes; other; none) | Help clinicians quickly flag visits that may be at higher risk for a diagnostic breakdown,[19,69] and to improve accuracy of the history by capturing symptoms in the patient or proxy's own words. | 6451 (91.2%) | 367 (79.8%) |
| | Medical history (free text) | Please tell us more about how you have been since your last visit | | 4724 (66.8%) | 264 (57.4%) |
| Getting it right (potential diagnostic process concerns) | Main concerns heard? (yes/no/have not discussed concerns yet ) | Thinking about the main symptoms or health concern(s) you want to discuss at your upcoming visit: Do you feel like your main health concerns (or what matters most to you) have been heard by your healthcare providers so far? | Enable early detection of PRDBs with the potential for high impact on the DxP[17] | 6664 (94.2%) | 407 (88.5%) |
| | Problems or delays with tests/referrals? (yes/no) | Have you had any problems or delays with tests or test results or referrals/appointments related to the health concerns you want to discuss at your visit? | | 6803 (96.2%) | 393 (85.4%) |
| | Other problems or delays? (yes/no) | Have you had any other problems or delays related to the health concerns you want to discuss at your visit? | | 6667 (94.2%) | 395 (85.9%) |
| | Description of problem or delay (free text) | Please tell us more about the most important problems or delays you noted above. | Details of PRDBs may facilitate response at the point of care, and offer a structured opportunity to address uncertainty in diagnosis[26] | 419 (57.0%)[a] | 96 (87.3%)[a] |
| | Description of what's working well (free text) | Is there something in particular that is going well for you in your care? | Appreciative inquiry: Identify things that are going well; feedback about helpful behaviors and experiences[70] | 1377 (19.5%) | 201 (43.7%) |

[a]Denominator restricted to those participants reporting a problem or delay.

Bell SK, Dong ZJ, Desroches CM, Hart N, Liu S, Mahon B, Ngo LH, Thomas EJ, Bourgeois F. Partnering with patients and families living with chronic conditions to coproduce diagnostic safety through OurDX: a previsit online engagement tool. J Am Med Inform Assoc. 2023 Mar 16;30(4):692-702. doi: 10.1093/jamia/ocad003. PMID: 36692204; PMCID: PMC10018262.

6. **Potential ADEPT implementation strategies**

Adapt and replicate AHRQ's The Toolkit for Engaging Patients to Improve Diagnostic Safety, which provides guidance on implementation e.g., Preparing you organization; Making a plan for implementation; Training your team.

Approach 1 (low intensity):
Team deploys the asynchronous Patient and Family Diagnostic Feedback Tool alone and use the AHRQ measure tools for evaluation.

Approach 2 (medium intensity):
Team deploys both the Patient and Family Diagnostic Feedback Tool and the SHARE Framework for Diagnostic Collaboration and monitors their use with the measurement strategies mentioned above.

Approach 3 (high intensity):
Scaling up, team could integrate this into medical learner and faculty development, involving a deliberate practice model.

7. **IHQSE training needs**

- A case for change will need to be created, as this level of patient/caregiver involvement may be challenging for frontline providers, who may perceive it as slowing down their workflow.
- An engagement plan for change management will be necessary.
- Leadership and stakeholder support will be needed to implement this intervention, particularly for educational, professional, and patient experience applications.
- Trainers and program evaluators will be required once the intervention is implemented and scaled.

8. **Assessment:**

**Cons:**
- May be perceived as time-consuming and may alter the typical 8-minute hospitalist visit (based on old time-flow studies).
- Interventions focused on physicians and does not at this stage includes other care team members e.g., nurses.
- Requires a culture change.

**Pros:**
- Achieves the quadruple aims:
- If it decreases diagnostic errors (DE) as expected, it will reduce costs across the population and improve overall health.
- Engages patients and families in their care and enhances the patient experience.

- Improves provider job satisfaction due to its patient-centered focus.

**Feasibility** – First two tiers are fairly feasible as they are fairly well operationalized as a questionnaire and scripted framework using a mnemonic.
**Impact** – Potentially high impact if done with rigor and leads to better metacognition and coproduction of diagnosis with patients/caregivers.
**Priority -** From the patient/caregiver's/societal perspective, likely very high.

### ADEPT Intervention Proposal

### Provider Self Reflection and Feedback

1. **Intervention descriptions:**

- Goals:
    - Improve diagnostic awareness, knowledge, attitudes, and skills for diagnostic excellence, related to cases seen by ADEPT reviewers as a primary aim, but as a lifelong clinical skill as a secondary aim
    - Use structured tools for clinicians to reflect on their performance in ADEPT cases
    - Provide direct resources in the form of coaching or clinical feedback if feasible
    - Share learnings generated by clinicians broadly

**<u>Principles and Key Components of intervention:</u>**

The group had strong agreement that structured self-reflection was key, with feedback on performance via sharing of our reviews or contact with ADEPT reviewers. Deploying this via survey-like tools (e.g., REDCap) would be feasible, with the expansion of support (e.g., coaching or provision of completed reviews) based on local capabilities.

**Self-Reflection component**:

a. Focus on self-reflection around events reviewed by ADEPT, using a structured format such as a modified SAFER Dx with guidance as to the concepts and practices the recipient should reflect on and guiding the recipient to consider some set of steps as they reflect on the case
b. Also provide information about what ADEPT has observed locally or overall.
c. Strong feeling that we should not have check-box Likert scale, but required fields that people should complete.
d. Apply this at least to cases we review, possibly also in broader range of cases we know take place but which are not selected at random.

**Feedback on performance in the context of Self Reflection**:

a. Use case reviews to share/give the information back to **provider** involved in a *beneficial way (Details provided later)*
b. Agreement that we share results with and without error

**Set community and cultural expectations for sharing self-reflection (No one learns alone)**

a. Add text box to reflection tool asking the person doing the reflection tool provide a teaching point back to their colleagues. Processing and sharing information from providers back to divisional community needs some additional thought.
    a. Processing text itself (redact, clean, remove PHI, etc).

b. All comments or some?
c. Individual or aggregate learning from self reflection tools
d. Can definitely share case review learnings back to providers

**Make this low impact on recipients**

- Limit the length and complexity of the review and deliver at a time (probably not while on service but also not long afterwards) where they can take some time to think.
- Attempt to maximize psychological safety – initial thoughts being, 1) wording of the reflection tool, 2) Buy-in and endorsement from Chief/ADEPT team, 3) at some sites, may need to ensure these are not discoverable and/or are protected from Peer Review activities.

**2. Priority ADEPT diagnostic gaps addressed with examples from our data:**

- Of the areas identified so far, key diagnostic steps which might be most amenable to self reflection and/or feedback-coaching include
  - Problems with Assessment
  - Followup and monitoring
  - Test selection and interpretation
- Other areas are less frequent but may also be useful:
  - History
  - Physical Exam

**3. Safety behavior target**

Safety I – Provider-facing

**4. Background evidence to support intervention:**

Like other patient safety issues, diagnostic performance is dependent on contextual factors such as workload, workforce, culture, and technology. But it is also a skill of individuals and teams of clinicians. If done in a psychologically safe way, self-reflection, feedback, and coaching regarding specific cases can improve clinicians' performance.

Enhancing physicians' diagnostic skills is crucial for improving patient outcomes and healthcare quality. Several strategies have been identified to facilitate this improvement, including reflective practices like Gibbs' Reflective Cycle, academic detailing, audit and feedback mechanisms, professionalism programs, and structured feedback after patient admissions or discharges. There are a few broad methods we might refer to:

**Gibbs' Reflective Cycle Applied to Physicians**

Gibbs' Reflective Cycle is a structured framework guiding individuals through a reflective process to learn systematically from experiences. It consists of six stages: Description, Feelings, Evaluation, Analysis, Conclusion, and Action Plan.
Implementing Gibbs' Reflective Cycle in medical practice can involve regular reflective writing exercises, facilitated group discussions, or integration into continuing medical education programs.

Reflective writing allows physicians to step back, review their thoughts, goals, and actions, and recognize how their perspectives impact their conduct. This process helps consolidate learning and boosts professional and personal development.   In general, this approach is felt to be best for adult learning but also hardest to sustain and most likely to be individual-dependent.

**Academic Detailing**

Academic detailing involves personalized education for clinicians, typically conducted through one-on-one sessions with trained healthcare professionals. The goal is to provide evidence-based information to improve clinical decision-making. Studies have shown that academic detailing can lead to improvements in targeted prescribing behaviors.

**Audit and Feedback**

Audit and feedback involve systematically reviewing clinical performance against established standards and providing feedback to clinicians. This approach has been associated with small to moderate improvements in professional practice, with effectiveness varying based on the context and delivery methods.

**Professionalism Programs**

Professionalism programs aim to enhance the professional behavior of clinicians, which indirectly supports diagnostic accuracy. Components may include workshops, mentorship, and reflective practices. Fostering a culture of professionalism is believed to contribute positively to clinical decision-making.

**Structured Feedback After events**

Providing structured feedback to physicians after events are thought to enhance diagnostic accuracy and overall patient care. There is intense interest in AI tools for this purpose.

**Physician Coaching**

Coaching is another approach which might be added to self-reflection to help guide improvement; coaching might be critical as part of this program as it may help improve diagnostic calibration in the learner/recipient. Principles of appreciative inquiry and improvisation *http://dx.doi.org/10.4300/JGME-D-19-00800.1* are thought to be useful in giving feedback around an individual case.

***Diagnostic excellence specific approaches:***

Self reflection:  Much of the work in this area has been done in the outpatient setting. The most widely promulgated tool is CalibrateDx: https://www.ahrq.gov/diagnostic-safety/tools/calibrate-dx.html (CalibrateDx Guide: https://www.ahrq.gov/sites/default/files/publications2/files/calibrate-dx-guide.pdf). CalibrateDx is not applicable to individual case reviews and was felt to be overlong.

Other models using self-reflection and self-evaluation exist (Rob El-Kareh's work, Trowbridge's work, and Gopi Astik's).  These tools have not always been tied to a clinical trigger event, but more to a handoff or a place in time where reflection is feasible. Few seem to have been sustained.

5. **References/Evidence:**

References available upon request

6. **Potential ADEPT implementation strategies**

**<u>Low intensity:</u>**

- Self-reflection on trigger events from ADEPT and those referred by Division
- Would require
    - Awareness campaign/coalition, and buy-in from Division (probably not Health System)
    - Educational campaign or toolkit to teach individuals about how to approach self-reflection
- Deployed via confidential REDcap survey, with reminder approach TBD.
    - Someone will need to add emails and MRNs to a new REDCap database (could use existing data pulls), as well as someone to manage questions as they come in.
    - 'No one learns alone' component also potentially addable here, with caveats as above.

**<u>Moderate Intensity:</u>**

- Low intensity model + providing results of completed reviews back to provider after asking provider if they want to have the "answer."
- Would require training and some logistical support for making sure we have a way to get our case reviews back to the provider in question.

**<u>Higher intensity:</u>**

Moderate intensity model with either direct coaching around reviews OR co-production of a review (e.g. correction or fill in gaps) with review team. The latter would be useful to ADEPT for improving our ability to understand context in complex cases but would probably also require that our team be aware of/be willing to push back on situations where we feel a diagnostic opportunity actually was present and the clinician disagrees.

Potential content modifications to CalibrateDX/SAFER

- Align with case review information wherever possible
- Make more about decompensation and recognition of common clinical scenarios when available
- Link to other supportive tools (EHR use tips, information on most common causes of errors overall, patient communication tips)
- Would need shortening, user pretesting before dissemination.

Other programmatic needs:

- Platform for review/reflection distribution (REDCap would be fine)
- Teaching and outreach materials (for wards or teams or divisions)
- Training or resources for any coaching or direct feedback if performed by a physician

**Potential Measurement Strategies:**

- Track number of conferences where materials distributed
- Performance of self-assessments by REDCap completion rates
- Track cases where they provided feedback
- Clinician surveys could also track satisfaction and any lessons learned, using a heavily modified Debrief Tool as a guide

| | Approach 1 | Approach 2 | Approach 3 |
|---|---|---|---|
| Plan | Create awareness campaign including educational sessions for providers | Get buy-in from clinicians regarding feedback; prepare site leads to give feedback | Get buy-in from clinicians regarding coaching; prepare site leads to coach |
| Do | Automatic email of tool to provider after triggers, data on DE rates, links to the self-assessment tools | Quickly adjudicate requested cases, provide data from case review back to primary provider, possibly team | Conduct coaching sessions (have to decide who would receive them) |
| Study | Follow-up surveys of providers in use of self-assessment, lessons learned | Site leads to track provision of feedback, response | Site leads to track provision of coaching, response |
| Act | Repeat messaging as needed | Iterative refinement based on above | Same |

7. **IHQSE training needs**
   Guiding coalition
   Alignment with business case
   Sense of urgency

8. **Assessment:**

Cons:

- Success will depend on receptivity of attendings and teams to feedback (may be easier with junior faculty or on teaching teams)

- Self-reflection quality will likely vary based on interest and skills of the attending unless tool is structured carefully
- Peer feedback and coaching requires additional time, which clinicians and teams may not have (for some divisions, individuals available for coaching may also be low

Pros:

- This may be one of the more potent interventions we do, both for individual cases and for changing culture

**Feasibility** – Moderate to high
**Impact** – Moderate
**Priority -** Moderate to high

### Provider Intervention:

### EHR training and best practices

1. **Intervention description**

- Materials to train clinicians in best practices using Epic to facilitate accurate and timely diagnoses and reduce EHR burden

2. **Priority ADEPT diagnostic gaps addressed with examples from our data:**

- Assessment
- Follow-up and monitoring
- Test selection and interpretation

3. **Safety behavior target**

Safety II – resilience (provider)

4. **Background and description of the intervention:**

OHSU (Gold, et al.) have been studying best practices in using Epic. These include training workshops aimed at residents (e.g., how to pre-round using Epic) and aimed at attendings (e.g., how best to trend data or find outpatient specialty provider notes). These are accompanied by a library of brief (3-8-minute long) videos on specific topics that users can access asynchronously as they wish. This intervention would be adapted for ADEPT.

Interested ADEPT sites that use Epic would receive:

1. Train the trainer workshops
    a. Who: Teachers: Jeff Gold and OHSU Clinical Informatics fellows. Attendees: ADEPT site leads, clinician Epic super-users, chief residents. They then train their own clinicians (residents, APPs, hospitalists) at each site.
    b. What: 90-minute session, aimed primarily at direct care providers (residents, APPs, hospitalists who provide direct care), with reference to attendings in supervisory roles (e.g., on teaching services).
    c. Where: virtual
    d. When: to be scheduled, may need more than one session so that everyone can attend. Sessions will be recorded for those who miss them.
    e. How: Jeff Gold and his clinical informatics fellows will slowly review each topic, attendees will have the opportunity to try out each feature in their site's Epic build, confirm it is there and if any modifications needed, ask questions, and learn how best to teach the material to their own clinicians.

    f. Why: this will help providers learn how to use Epic most effectively and efficiently, enabling them to gather data necessary to make correct diagnoses, monitor for failure to respond as expected, and identify potential complications. It may also free up clinician time to think, talk to patients and caregivers, and take other steps to enable a best possible diagnostic process.
2. Videos of workflows and simulations Jeff Gold has created (and may create more)
    a. Who: ADEPT site leads, clinical informatics fellows, clinician Epic super-users, and clinicians at each site
    b. What: library of short online videos regarding various topics in using Epic, all aimed at inpatient medicine clinicians involved in the diagnostic process
    c. Where: virtual
    d. When: on-demand
    e. How: ADEPT site leads +/- CI fellows and Epic super-users will first preview each video and make sure it is applicable to their site's Epic build. If it is, they will be made accessible to residents, APPs, and attendings on medicine, along with an online user's guide (table of contents with links, differences between Epic in video and local build). Videos that are not applicable will be deleted.
    f. Why: Epic has many ways to do each task, but only a few are optimal. By demonstrating the most effective and efficient ways to do each task, clinicians will have better diagnostic performance for all the same reasons noted above.

Measurement:

- Site lead attendance at train the trainer sessions
- Number of attendees at each site's training sessions
- Number of downloads of each video
- Use of "high-yield" Epic features using Signal data, slicer-dicer data model, reporting workbench, or custom reports
- Audit log data
- DE rates once implemented, distribution of contributing causes to DE, severity, amount of delay
- Provider survey (pre-post) – do you use these features? How valuable are they?
- Number of sites that choose to implement
- At each site, the number of best practice tools that are present in their build
- For adjudicators: does time for adjudication go down?

#### 5. References/Evidence:

During the COVID pandemic, when there was a need for rapid deployment of clinicians to the inpatient setting, including the ICU, clinical informatics fellows and subject matter experts at OHSU developed and deployed a tailored EHR curriculum comprising of in-person classes and online video modules, leveraging high-fidelity simulated patient cases, similar to what is proposed here.[1] All recipients of the training (nurse anesthetists being deployed to the ICU) felt the training was useful and improved their EHR skills, with a Net Promoter score of +87. The training increased comfort in the ICU for all respondents, and 91% felt the training prepared them to work in the ICU with minimal guidance. 58% viewed the online video library.

Other research in this area has involved the use of an EHR simulation environment using cases with embedded safety issues that clinicians needed to find. In one study, residents were given an immediate debriefing regarding missed issues and strategies for data gathering in the EHR. Repeat testing with a different case 1 week later demonstrated improvement in individual performance from 39.9% to 63.6%, including higher rate of recognition of changes in vitals, misdosing of antibiotics, and oversedation compared with first-time participants.[2] Simulation-based EHR training may also reduce physician burnout by optimizing their efficiency.[3]

#### 6. Potential ADEPT implementation strategies

Approach 1 (low intensity):
Train the trainer, presentations at division meetings and residency meetings, give access to curated set of videos, customized guide by site

Approach 2 (medium intensity):
As in Approach 1, also provide videos as part of starting on service orientation email, take more steps to make sure everyone is exposed to these videos and training sessions

Approach 3 (high intensity):
As in Approach 2, also provide 1:1 coaching, especially if evidence of suboptimal use of Epic based on Signal data (or Jeff Gold's custom reports). Deliberate practice with real patients. If site has capability, could also create simulated cases with issues that need to be identified; create custom videos for site and make part of required annual training.

| | Approach 1 | Approach 2 | Approach 3 |
|---|---|---|---|
| Plan | Train the trainer; customize video guide for each site | As in 1, plus do more of a campaign, plan orientation sessions. Pre-survey. | As in 2, plus prepare to get Signal, slicer-dicer, reporting workbench, and/or custom reports. Customize videos for site. Create simulated cases. |
| Do | Give presentations to attendings, APPs, and residents; give access to videos | As in 1, plus provide videos as part of orientation to rotations | As in 2, but with custom videos. |
| Study | Track attendance at meetings, video downloads; get informal feedback; track DE rate data | As in 1, plus reach out to those who didn't attend, get them to attend | As in 2 plus use Signal and Report data to track individual use of best practices |

| | | | |
|---|---|---|---|
| | | additional sessions. Post-survey. | |
| Act | Give additional presentations, revise video guide as needed | As in 1, plus integrate survey data. | As in 2, plus 1:1 coaching if evidence of suboptimal use of best practices. Case simulation or deliberate practice. |

7. **IHQSE training needs**

Guiding coalition

May need to add informatics fellows and Epic super-users, chief residents, whoever is in charge of APP and attending onboarding.

Alignment with business case

Has potential to be low-cost/high-yield

Sense of urgency

Link back to underlying causes, esp. chart biopsy, monitoring.

8. **Assessment:**

Cons:

- Most potent versions of the intervention likely require site customization (videos, simulated cases) and/or more intensive implementation (reports, coaching)
- Users who struggle the most may have trouble with videos that are not customized for their site, even with a guide

Pros:

- Feasible, but effectiveness likely depends on intensity of implementation
- Easy to measure use of videos, attendance at training sessions
- Could be monitored with Signal data, custom reports, etc.

**Feasibility** – Moderate
**Impact** – Moderate
**Priority -** Moderate

### ADEPT Intervention Proposal

### Diagnostic Time-Out (DTO)

1. **Intervention description:**

Goals:

Aim to integrate a structured, evidence-based diagnostic reassessment (Diagnostic “time-out”) tool into Hospital Medicine teams' workflow, prompting clinicians to systematically reconsider the working diagnosis in real time. We aim to create a tool for use both in the educational setting (on teaching service teams) as well as for independent direct-care providers.

**Key Components of a Diagnostic Time-Out Tool:**

Based on a review of the literature and existing DTO programs at other institutions, two key components must be in place for successful implementation

- A structured step-by-step DTO process, commonly with a tool to document the outcomes of the diagnostic pause
- A shared set of triggers (including patient, provider, and system factors) to initiate a DTO

**Diagnostic Time-out process**
Current iterations of DTO tools vary somewhat in their components. For ease of use and widespread uptake, we outlined the following DTO structure

1. **Identify the working Diagnosis**
2. **Identify what data does and doesn’t fit** with the working diagnosis
    - Consider specific cognitive debiasing prompts
        - *Have we anchored on a diagnosis prematurely?*
        - *What findings don’t fit our initial impression?”*
        - *Are we ignoring any data because it contradicts our favored diagnosis?*
        - *Could this be an atypical presentation of a common illness?*
3. **Discuss alternative diagnoses**
    - Consider formally revising/reprioritizing the differential diagnosis
4. **Plan for next steps –** be specific about changes to the diagnostic/management strategy
    - *Are there additional consultants to call?*
    - *Any new diagnostic studies?*
    - *Access additional data?*
    - *When to reassess the working diagnosis?*

**Diagnostic Time-out Trigger** - We suggest considering a structured DTO in any of the following scenarios

- Any time a high degree of uncertainty is present
- If undifferentiated symptoms remain >24h after admission
- During transitions of care, including:
  - Inter-service transfers
  - Transfer from an outside hospital
  - Night team to day team handoff
- **Escalations of care, including:**
  - Rapid Response call
  - Transfer from acute care to step-down unit
  - Transfer to the intensive care unit (ICU)
- If clinical course is not as expected
- If limited history/physical findings are present (AMS, language barriers, isolation precautions)
- Multiple consulting services involved
- When any team member has concerns about the diagnosis
- If there is a discrepancy between patient and provider understanding of the diagnosis

2. **Priority ADEPT diagnostic gaps addressed with examples from our data:**

- Of the areas identified so far, key diagnostic steps which might be most sensitive to DTO
  - Problems with Assessment
  - Followup and monitoring
  - Test selection and interpretation
- Other areas are less frequent but may also be useful:
  - History
  - Physical Exam

3. **Safety behavior target**

Safety II – Provider-facing

4. **Background and description of the intervention:**

The Diagnostic Time-Out (DTO) is an established strategy for addressing diagnostic errors, with several studies supporting its use. Based on existing patient safety strategies, such as the surgical time-out, the DTO applies cognitive debiasing techniques to counteract common errors in diagnosis. It is a structured pause in clinical reasoning that prompts clinicians to systematically reassess the working diagnosis.

DTO effectiveness has been measured across various healthcare settings. Several pediatric hospital medicine groups have implemented DTOs, with evidence for both feasibility of use as well as positive diagnostic outcomes. In the intensive care unit (ICU), the ICU-PAUSE tool integrates a diagnostic reassessment step during handoffs, ensuring continuity of diagnostic reasoning and preventing diagnostic momentum. Similarly, in emergency medicine, brief DTO checklist caused clinicians to broaden their differential diagnosis and have been associated with an increase in diagnostic accuracy. Additionally, the CDC recommends the implementation of a diagnostic pause as a component of their “Core Elements of Hospital Diagnostic Excellence.”


#### 6. Potential ADEPT implementation strategies

The precise DTO intervention will likely vary by institution. We suggestion adoption of our ADEPT DTO toolkit for success in both implementation and measurement of the DTO intervention

**ADEPT Diagnostic Time-Out Toolkit will include**

- *Pocket card / web app detailing the DTO process and suggested triggers*
- *REDCap survey for tracking cases and collecting use/outcome data*
- *EHR Dot Phrase for documenting clinical reasoning (and collecting use data)*
- *“Reach Suggestions” for select institutions who wish to implement more structured diagnostic time-out programs (this could include an educational program, interdisciplinary DTO huddle, cultural expectation to document DTO in EHR, etc)*

***Low Intensity*** *– not recommended*
- *Distribute toolkit*
- *Awareness campaign (i.e. posters with QR code)*

**Medium Intensity**
- Cultural expectation of use
- Educational intervention (i.e., residency campaign)

**High Intensity**
- Specific DTO program with interdisciplinary team
- Specific educational program
- EHR documentation expectation

**Examples from existing programs**

- Vanderbilt: overnight Hospital Medicine team at the VA hospital performs a DTO every night (1 intern selects a patient for DTO each shift). DTO used as an educational tool as well as quality improvement intervention.
- Children's Hospital of Colorado developed DTO toolkit including a detailed clinical pathway algorithm

**Measurement Strategies**

We suggest a tiered measurement strategy of the Diagnostic Time-Out tool

- For all sites, the intervention team will measure DTO tool uptake using one or more of the following:
    - Survey of faculty and residents on the use of the DTO tool, its effectiveness, barriers to intervention
    - Direct measurement of update through a REDCap DTO tool
    - Tracking use of an established "DTO dotphrase" in sites' EHR
- As a secondary strategy at certain sites, the intervention team will measure targeted diagnostic outcomes:
    - Patients undergoing DTO will be identified using REDCap survey
    - Retrospective review of cases could include identification of changes to the working diagnosis, broadening of differential diagnosis, changes to management strategy, etc.
    - Understanding of patients/diagnoses most likely to require DTO can inform further interventions

7. **IHQSE training needs**

   Guiding coalition
   Alignment with business case
   Sense of urgency

8. **Assessment:**

Cons:

- Largely educational intervention, limited evidence for how to implement effectively in multiple settings

Pros:

- Has alignment (conceptually) with case review process in that it covers key steps in diagnosis
- Could be enhanced with concepts like prospective hindsight, or baked into handoffs or RRT debriefs

**Feasibility** – Moderate to high
**Impact** – Low Mod
**Priority -** Low Mod

### ADEPT Intervention Proposal

#### Embedding Diagnostic Safety into Safety and Quality infrastructure

1. **Intervention descriptions:**

Goals:

- Build diagnostic error (DE) review program into existing patient safety systems at each hospital so that patient cases can drive and inform system-level improvements.
- Make this program sustainable beyond the ADEPT grant time period.

**Principles and Key Components of intervention:**

- Leverage ADEPT teams to form a hub-and spoke model, where a division- or department-based Diagnostic Excellence team (or a steering committee or similar program) shares information about case findings with the Department of Quality and Safety (or similar entity).
    - ADEPT/hospital medicine team members will provide expertise in creating case review infrastructure and training adjudicators for other divisions/departments (as and when they decide to participate in the program).
    - All divisions/departments will handle their initial reviews; they will share findings/narratives locally/within department and escalate trends/system-level issues to the hub.
    - *(Rationale for creating separate review teams by division/department: clinical experts from respective departments are best suited to identify DEs, opportunities for improvement, and plan targeted interventions)*
- Identify and engage key stakeholders (quality and safety team, clinical leadership, residency program leadership, etc.)
    - Use strategies learned at IHQSE.
- Define specific roles, create committees, and provide appropriate training (e.g., adjudicators, committee to analyze cases/trends and give recommendations).
- Ensure peer review confidentiality is maintained; get processes reviewed by risk/legal teams. Need to have a secure, confidential database; explore options for a local version of REDCap or other database solutions.

2. **Priority ADEPT diagnostic gaps addressed with examples from our data:**

Likely addresses multiple, though may initially focus on systems-based problems more than provider performance

3. **Safety behavior target**

Safety I – System

4. **Background and description of the intervention:**

**Background:**

- One of the primary goals of the ADEPT study is to incorporate surveillance for diagnostic errors into usual care at each hospital.
- Based on our learning from the ADEPT study, diagnostic opportunity events are often not detected through traditional incident reporting systems.
- Cases of DE (+/- harm) can be submitted for formal case review using the hospital's usual incident (quality/safety) reporting system and depending on available resources/infrastructure, can additionally be analyzed as part of a Diagnostic Excellence (Dx Ex) or similar program.
- This will lead to the identification of system-level contributors to DE that can then serve as target for interventions.
- This is a base requirement for any effort to improve diagnostic performance at the system level. Sustaining this effort may require commitment or resources beyond ADEPT study duration.

Approach 1 (low intensity):

- Cases of DE (+/- harm) submitted through existing trigger events and/or safety event reporting system.

    *(Rationale for using the existing system: maximum feasibility and sustainability)*

Approach 2 (medium intensity):

- Approach 1 plus 10 randomly selected cases of ICU transfers and inpatient deaths every month.

    *(Rationale for excluding RRTs: criteria for calling RRTs vary widely across sites; ICU transfers and deaths are associated with maximum harm and are, therefore, of greatest clinical interest)*

Approach 3 (high intensity):

- Approach 2 plus one of more of the following:
    - Additional randomly selected cases of ICU transfers, inpatient deaths.
    - Random sample of rapid response events, early readmissions.
    - Other EHR based triggers: existing early warning systems, deterioration indices.
    - Clinician reported events/feedback (in addition to existing general safety event reporting system). Examples include:
        - Mandatory mortality reviews (or similar surveys sent to clinicians after all cases of ICU transfers, RRTs, etc.).
        - Clinician surveys sent for each randomly selected case identified for review (ADEPT approach).
        - Create a parallel reporting system dedicated only to diagnostic safety events.

- Patient reported events/experience surveys (with involvement of PFAC).

5. **References/Evidence:**

   Diagnostic excellence specific:

   Measure Dx: A Resource To Identify, Analyze, and Learn From Diagnostic Safety Events. Content last reviewed March 2024. Agency for Healthcare Research and Quality, Rockville, MD. https://www.ahrq.gov/diagnostic-safety/tools/measure-dx.html
   Graber ML, Castro GM, Danforth M, et al. Root cause analysis of cases involving diagnosis. Diagnosis (Berl). 2024;11(4):353-368. Published 2024 Sep 3. doi:10.1515/dx-2024-0102

   Obviously a broader literature exists to describe safety programs and best practices.

6. **Potential ADEPT implementation strategies**

**Step 1. Case Identification:**

- Approach:
  - Identify an enriched cohort of cases with enough diagnostic errors and process faults/missed opportunities to learn from.
  - Use caution in adding additional workload for sites – balance sensitivity and specificity of case identification.
  - Need for tiered approach – based on institution buy-in and available resources.
- Tier 1 (low intensity):
  - Cases of DE (+/- harm) submitted through existing safety event reporting system. *(Rationale for using the existing system: maximum feasibility and sustainability)*
- Tier 2 (medium intensity):
  - Approach 1 plus 10 randomly selected cases of ICU transfers and inpatient deaths every month.
    *(Rationale for excluding RRTs: criteria for calling RRTs vary widely across sites; ICU transfers and deaths are associated with maximum harm and are, therefore, of greatest clinical interest)*
- Tier 3 (high intensity):
  - Approach 2 plus one of more of the following:
    - Additional randomly selected cases of ICU transfers, inpatient deaths.
    - Random sample of rapid response events, early readmissions.
    - Other EHR based triggers: existing early warning systems, deterioration indices
    - Clinician reported events/feedback (in addition to existing general safety event reporting system). Examples include:
      - Mandatory mortality reviews (or similar surveys sent to clinicians after all cases of ICU transfers, RRTs, etc.).

        - Clinician surveys sent for each randomly selected case identified for review (ADEPT approach).
        - Create a parallel reporting system dedicated only to diagnostic safety events.
    - Patient reported events/experience surveys (with involvement of PFAC).

- All ADEPT sites should aim for either Tier 2 (medium intensity) or Tier 3 (high intensity) case identification approach based on available resources.

**Step 2. Case Review:**

- Create a simplified, efficient, and reliable case review process.
    - Use structured, validated tools:
        - Safer Dx (to accurately assess diagnostic errors)
        - DEER taxonomy (to identify diagnostic process failure points)
        - NCC-MERP (to categorize the degree of harm among cases with DE)
    - Two-tiered review process: rapid single adjudicator (MD/APP) review for a larger number of cases and in-depth 2-clinician/expert group review for selected cases.

  *(Rationale for a two-tiered review process: ADEPT case reviews so far have demonstrated strong inter-rater reliability; a two-tiered review process will help maximize efficiency and reliability)*
- New case adjudicators can be trained using a case from the existing training materials created by the core ADEPT team and then will participate in experiential learning. Would ideally have some experience in pair/group prior to serving as rapid single-adjudicator.

**Step 3. Measure and Analyze:**

- Quantitative and qualitative analyses to look for trends and identify opportunities for improvement/interventions.
    - Use Root Cause Analysis (or mini-RCAs, fishbone diagrams) for qualitative analysis and to generate narratives to guide interventions. Consider adapting the RCA structure to evaluate cases involving diagnostic error, including a focus on the cognitive aspects of medical decision-making
    - Use Healthcare Failure Mode and Effects Analysis (HFMEA) to systematically identify and address failure points.
    - Need to track demographic factors to ensure equity in diagnostic safety initiatives.
- Currently the goal is to flesh out rationale for each intervention with narratives from series of case reviews. In future, can aim to develop standardized measures (such as DE rates) that can be used to benchmark performance over time and across sites.

  *(Rationale for avoiding focus on DE rates at this time: there is wide variability in patient populations and in the criteria for RRTs and ICU transfers across sites, making it challenging to compare rates across sites; there may not be enough cases of DE at individual sites to reliably measure changes in DE rates over time)*

**Step 4. Act:**

- Align the frequency of data sharing with specific goals.
- Establish a database of lessons and interventions resulting from diagnostic case reviews

- Need for integrating the infrastructure used to measure and analyze diagnostic errors and process failures with ongoing improvement initiatives, utilizing PDSA cycles.

- Within Division safety team:
  - Monthly cadence of case review, discussion, action item dissemination
  - Develop a business case to support diagnostic excellence initiatives and present this to key leaders (CQO, etc.)
- Within larger division/department:
  - Share cases with peers and trainees (grand rounds, noon conferences, M&M, etc.).
  - Create a diagnostic excellence curriculum (including a database for lessons learned from exemplar cases)
  - Create a shout out/celebration program (Dx Flex).
  - Share trends and aggregate results with division/department leadership on a quarterly basis, including a dashboard highlighting diagnostic error rates from case reviews
  - Identify opportunities for improvement/interventions.

- Department of Quality and Safety:
  - Submit cases into the existing incident reporting systems on an ongoing basis.
  - Share trends and aggregate results with Quality and Safety leadership on a quarterly basis, including a dashboard highlighting diagnostic error rates from case reviews
  - Identify system-level opportunities for improvement/interventions.

*Potential Measurement Strategies*:

- Consider the RE-AIM framework:
  - Reach: Proportion of teams or units where the review of DE is built into usual care.
  - Effectiveness: Number of cases reviewed; number of DE and process faults identified.
  - Adoption: Number of DE cases analyzed quantitatively or qualitatively to identify opportunities for improvement.
  - Implementation: Number of Safety I and Safety II interventions rolled out as a result of the case review process.
  - Maintenance: Track the above metrics over time to show sustained effectiveness.

#### 7. IHQSE training needs

Sense of urgency
Managing up
Voice of the customer

8. **Assessment:**

Cons:

- Case reviews take time and are hard to sustain
- Safety groups at our hospitals already fairly stretched
- No clear path towards provider performance improvement from health system (tends to trigger peer review)

Pros:

- Fits within existing frameworks well
- Many sites already reaching out to ADEPT teams to do additional reviews outside of our research

**Feasibility** – Moderate
**Impact** – Moderate
**Priority –** Required as part of ADEPT study

### ADEPT Intervention Proposal

### Second Opinion Programs to Improve Diagnostic Performance

**Second Opinion**

1. **Intervention Description**

Although clinical questions frequently arise related to diagnostic reasoning, there is a dearth of formal programming to facilitate communication about clinical conundrums as part of a Second Opinion model to support diagnostic processes.

Cases that may benefit from a Second Opinion consultation include 1) Those with diagnostic and/or therapeutic uncertainty, with a clinical query that does not fall under the umbrella of a particular subspecialty consultation, in whom such subspecialty consults have been unable to adequately guide therapy, or for which the question is logistical or ethical in nature; and 2) Cases which require real-time evaluation, such as a patient having a rapid response, e.g. assistance with assessing a patient's volume status and best next steps in therapy.

**Key Components of Consult Service**

Based on our review of the literature and experience as clinicians and ADEPT reviewers, we identified the following as key facets of the structure of a Second Opinion consult service:

- Real-time case review: Having someone available to review the case in real-time, particularly at times when staffing may be thin
- Consultant at one's own institution to facilitate review of the primary data via Electronic Medical Record
- Growth mindset that destigmatizes and normalizes asking for help

2. **Priority ADEPT Diagnostic Gaps addressed:**

Assessment, Teamwork, Communication

3. **Safety Behavior Target**

Safety II – Provider and System

4. **Background and literature to support the intervention**

We categorized the extant frameworks to consider for extrapolation to Second Opinion consultations as follows:

- Tumor board model/local collective intelligence: Scheduled multidisciplinary meetings to guide therapeutic decision-making at broad chronologic intervals, centering around an oncologic diagnosis.
- Single-clinician consultation model: User satisfaction and impact on referring clinicians' course of action has been studied; however, no diagnostics-related outcomes have been examined. Second opinion studies, in which a previously made diagnosis is re-examined by another expert, primarily center on oncology-related diagnoses in the fields of radiology and pathology. They focused on diagnostic and therapeutic consistency. A couple of studies looked at eConsults from primary care providers to specialists, including general medicine specialists; PCPs felt they had received good advice that either changed or confirmed their course of action.
    - Virtual consult (e.g., eConsult)
    - Senior physician as sounding board
    - Near-peer as consultant
- Virtual collective intelligence: Virtual platforms that allow posting of diagnostic/therapeutic questions with multiple user response. These vary in the number or expertise of user, the median response time and the type of questions posed. Questions trended toward therapeutic questions. In the medical diagnostic realm, users were most likely to inquire about diagnostic test characteristics.
    - Mobile Application, DocCHIRP: Hospital-medicine focused, 85 users, 45 consult questions over 244-days, median response 19min, themes around complex medical decision making and inquires re: prescription medicine use (Sims et al, JHM)

5. **References**

See above

6. **Potential implementation strategies:**

**Consult Structure**

The structure of the consult service would likely vary to some degree depending on existing structures. We would categorize or label potential consult models as 1) those that require extra resources to set up; and 2) those that do not require additional resources. The following potential interventions will serve as scaffolding for institutions to modify according to their clinicians' needs:

- Lower-tier interventions
    - Dyads or pods between attendings on service for first-pass discussions (a clinician's "Diagnostic Pod"), particularly if physical proximity of attendings already exists, such as in a large work room, and ensuring nocturnists are paired with other nocturnists to facilitate on-the-fly discussion.
    - Emphasizing a goal to practice in a common work room to facilitate discussion.
- Higher-tier interventions

- A Second Opinion clinician of the day who would contact clinicians in the morning, normalizing uncertainty and offering to assist with discussion of the case
  - This individual could sign into a created service line "Second Opinion- Hosp Med" and theoretically receive consults or SecureChats
- An on-call pool ("Diagnostic Management Team") for further discussion that can be contacted via email or secured messaging (e.g., Voalte)
- Secure LLM Consult to establish access to secure, enterprise LLM for use with PHI
  - At minimum, one member of the institution's Second Opinion team would be an expert in the current technology available and institutional policies around the use of LLMs for clinical queries
  - Optimally, the Second Opinion team would also disseminate best practices regarding LLM use for second opinions

Measurement approaches: We would measure frequency of use via brief REDcap form to track consults. Our measurement strategy would focus on:

- The provider experience and satisfaction, centering on burnout and isolation
- The consulting clinician's perception of how a Second Opinion consultation changed management
  - Broadening the differential such as by preventing misdiagnosis or leading to involvement of a subspecialty consultant
  - Changing diagnostic approach
- Keeping track of cases discussed, including diagnoses and areas of uncertainty, for use in educational conferences and materials.

7. **IHQSE training needs**

   Guiding coalition
   Measurement strategy
   Business case

8. **Assessment**

Cons:

- Although there is an evidence base for collective intelligence in medical diagnosis, it is unclear if our proposed dyadic model leads to diagnostic process or outcome change.
- Measurement by survey leads to additional provider burden and risk for low response rates.

Pros:

- On-service dyadic or pod model would be resource neutral though would require training and some mechanism to ensure the model is utilized.
- Not all providers have similar psychological safety with one another.
